# Parental socioeconomic position, childhood poverty and healthy ageing: retrospective and prospective study of six cohort surveys across 32 wealthy and developing nations

**DOI:** 10.1101/2025.08.22.25334234

**Authors:** Gindo Tampubolon

## Abstract

The childhood poor in wealthy countries have reported worse cognitive, muscle and mental functions as well as more frailty and multimorbidity as older adults. But it is uncertain whether the childhood poor around the world fall short of attaining healthy ageing because information of childhood conditions is often erroneous. Here I present new evidence on the life course shaping of healthy ageing among older adults around the world.

**METHODS:** Some 80 thousand older adults over 50 years in 32 wealthy and developing nations recalled their childhood conditions at ten to fourteen, then prospectively reported their health functionings. By the decade’s end these countries host more than half (53%) of the world population of 60 year and over according to the United Nations. Following recent empirical studies, a modified healthy ageing scale is constructed using a generalised latent trait model. The childhood conditions in England, Ireland and continental Europe include numbers of books, rooms and people (indicating overcrowding), presence of running hot water and central heating. Across in America, these are mostly replaced with financial hardship or family indebtedness; in China starvation to death due to government edict while in Indonesia presence of running cold water. Per prior practice childhood poverty is a latent construct of these error-laced recollections. Its associations with healthy ageing scale (modified) are obtained with fixed effects model, controlling for parental socioeconomic position, age, sex, education, wealth and marital status. Extensive sensitivity analyses assessed robustness.

**RESULTS and DISCUSSION:** Childhood poverty is associated with reduced healthy ageing by 0.14 standard deviation (z = -17.6); older adults who grew up poor have lower healthy ageing scores relative to those who were not poor. And women reported lower healthy ageing scores. Distributions of healthy ageing vary across countries, as do age profiles of healthy ageing in older adults. Childhood poverty is common in developing countries and non-negligible in rich countries. Social and economic progress over the long peaceful century is no guarantee of a complete poverty eradication promised by the world leaders in the UN 2030 Agenda for Sustainable Development. In fact, lifting all children out of poverty is set to face more challenges with regional wars in across the world. Childhood recollections show that the childhood poor grow old fall short of healthy ageing, making the life course shaping of healthy ageing central. And the strong evidence presented here calls for urgent actions to eliminate child poverty on account of its lifelong rewards. Researchers can now use childhood recollections across wealthy and developing nations alike, despite their measurement errors and their variation in elicitation, to estimate their associations with older adults’ health. And health policy makers should adopt a life course approach when evaluating childhood health interventions on account of their lifelong associations.

## INTRODUCTION

The 21st century opened with an unprecedented demographic shift: population ageing is accelerating worldwide, presenting ongoing challenges to society, welfare states and healthcare systems. The scale and speed of ageing in developing nations pose a concentrated challenge, what took France a century will take China merely a few decades, according to the World Health Organization.^1^ The United Nations has thus designated 2021–2030 as the UN Decade of Healthy Ageing, signalling a global commitment to foster longer lives that are healthier.^2,3^ This requires a fundamental re-evaluation of how health is promoted across the human life course. Often measures of health have been limited to specific populations and emphasised the absence of disease. Fortunately, a more fruitful understanding of health in the context of population ageing has emerged. The World Health Organization (WHO) defines healthy ageing as "the process of developing and maintaining the functional ability that enables wellbeing in older age".^1,4^ This functional ability is dynamically shaped by the interaction between a personal intrinsic capacity and their environment, the person’s physical and mental faculties such as cognition, psychological state, vitality, locomotion, and sensory functions.^1,5^

Empirical research on healthy ageing has mainly followed two approaches as exemplified by two studies in 2021, the healthy ageing scale^6^ (implementing a latent trait approach) and the healthy ageing index^7^ (implementing a counting approach).^8^ While the approaches differ, both the resulting constructs aim to capture the biopsychosocial aspects of health and functioning, integrating domains of vitality, sensory functions, locomotion, cognition, and activities of daily living that reflect an individual’s interaction with their environment (though the healthy ageing scale conspicuously omit mental health item in its construction).^6,7^ Such a multi-dimensional approach is vital because social engagement and mental capacities are as important as biological factors for achieving healthy ageing.^1–3,5,9^

### Two fife course frameworks

A critical instrument in explaining healthy ageing is the life course framework of health which has been receiving mounting empirical support.^10–17^ This posits that health outcomes are not exclusively determined by current conditions but are shaped by a complex interplay of protective and risk factors throughout life, from before birth to old age. This temporal and societal perspective emphasizes that investments in health at any life stage yield compounded benefits over time and for subsequent generations, reminding that factors in early periods can shape physical and mental capacities in later life including muscle function, cognitive function and mental health.^10,13,14^ Critical and sensitive periods (e.g., early childhood, adolescence, mid- life) and significant transition periods (e.g., entering school, parenthood, retirement) are recognized as pivotal moments where exposures can have profound and lasting effects on health trajectories. Here are two recent frameworks on the life course shaping of older adults’ health (first, dementia from the Lancet Commission and second, broader outcomes from the WHO)^5,18^. Comparing the two frameworks, the WHO framework appears more elaborate at the childhood stage and is used here to guide research in particular highlighting the role of overcrowding and housing quality. These specific features have been amply shown to shape older adults’ health in up to 31 wealthy and developing nations in single and cross-country studies, explaining gait speed, depression, episodic memory, probable sarcopenia, frailty and multimorbidity.^13–15,19^

**Figure 1.**
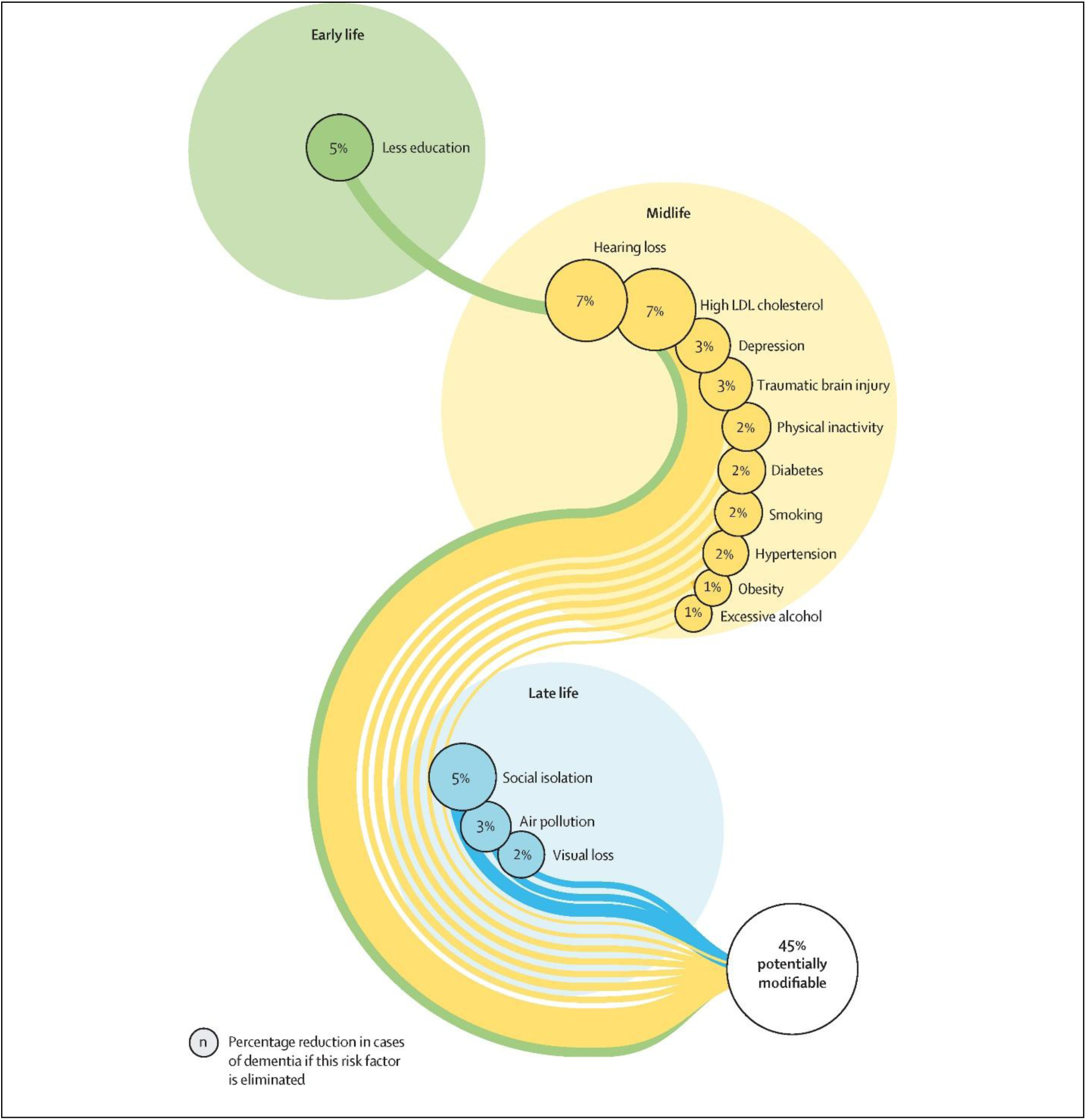
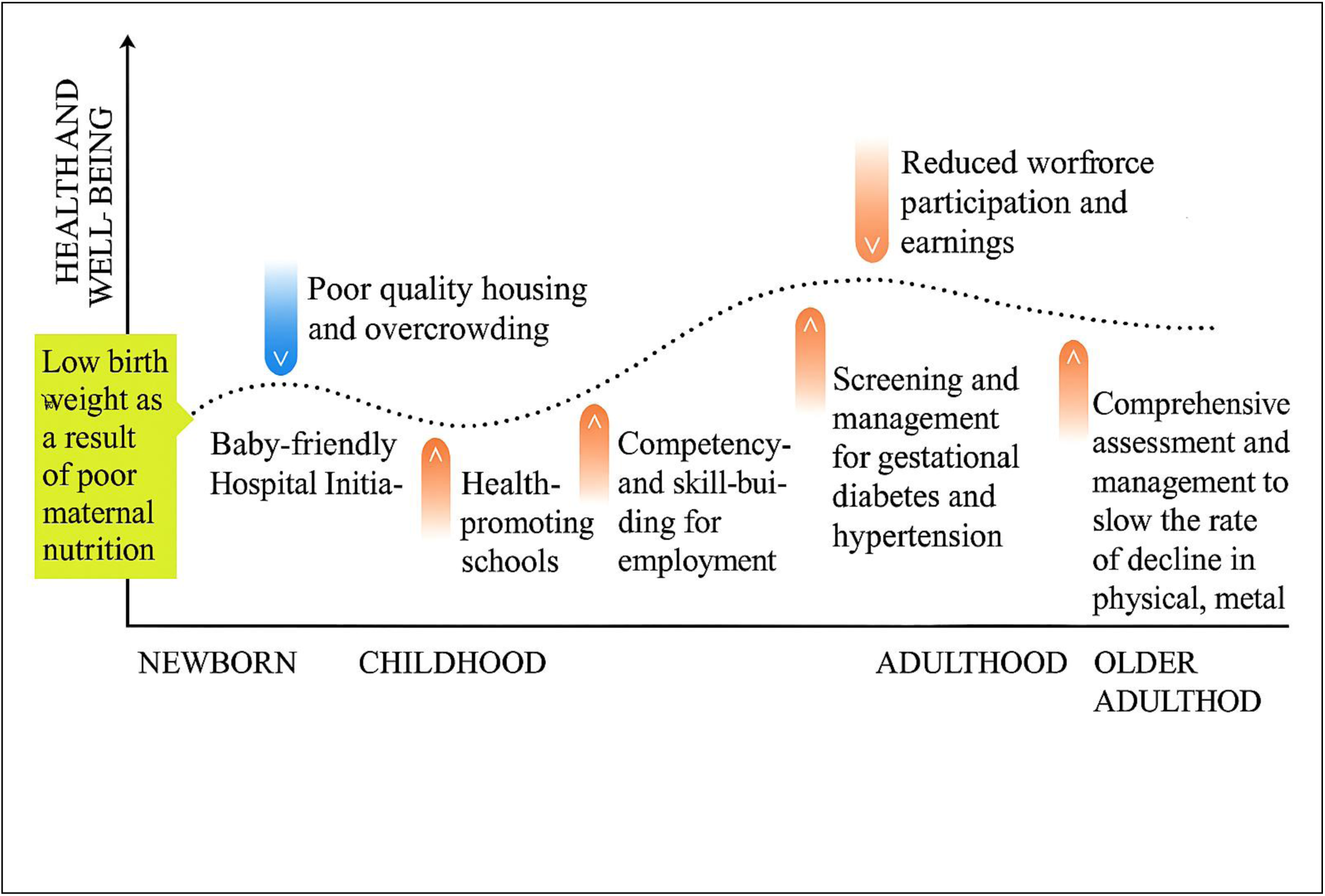
Two life course frameworks: top, Lancet Commission on Dementia (2024) and bottom, WHO Framework to implement a life course approach in practice (2025)^5,18^

Evidence has repeatedly shown that childhood poverty is a risk factor for old age disability, dysfunction and disease across wealthy and developing nations including the US, Britain, China and Europe. A recent study significantly expands this understanding, presenting the first analysis across 31 diverse countries to demonstrate that childhood poverty is a significant predictor of multimorbidity or multiple chronic conditions in later life.^19,20^ This association, with childhood poverty linked to a higher probability of multimorbidity, holds true across varied health systems, from the UK National Health Services to largely private systems in the US. The study draws on five major longitudinal cohorts – the US Health & Retirement Study (HRS), the English Longitudinal Study of Ageing (ELSA), the Survey of Health Ageing & Retirement in Europe (SHARE), the China Health and Retirement Longitudinal Study (CHARLS), and the Indonesia Family Life Survey (IFLS) – encompassing a broad spectrum of ageing experiences at different rungs of socioeconomic development. In the study I have suggested a potential epigenetic explanation with a supporting evidence from the US where childhood poverty is found to associate with more adverse epigenetic changes through increasing methylation rates captured in old age.^14,21^ This extensive geographic reach and the diversity health systems under examination suggest that the life course shaping of healthy ageing hypothesis can be usefully examined elsewhere.

Obtaining such evidence also served to highlight methodological challenges when applying the life course framework to explain later life health especially when the framework relies on retrospective information of the childhood stage several decades in the past. I found for instance,^13^

In 2008 Britons aged 50 were asked to recall the number of rooms and people in the house when they were 11, in order to assess overcrowding. Only one in three got both numbers right (compared to their mothers’ original answers when visited those years ago; both numbers are required to measure overcrowding).

The same cohort i.e. the 1958 National Child Development Study^22,23^ were again visited when aged 62 in 2020 - I found only 33.5% got both numbers right. Across the oceans, in Indonesia in 2015 I found fewer than one in ten older adults got both numbers right.^19^ Retrospective information of childhood conditions elicited from older adults living in wealthy and developing nations is mostly erroneous. Yet, most empirical studies use the information as error-free.

Fortunately, in recent studies, the use of a latent construct of childhood poverty is essential to handle erroneous recall when researching the life course shaping of health in one or many _nations._12–15,19

Beyond childhood poverty, broader socio-economic inequalities consistently influence healthy ageing across nations. Research comparing the US, England, China, and Japan shows that older people with disadvantaged socio-economic positions are less likely to achieve healthy ageing than those with advantaged positions".^7^ This multinational comparison, which also drew from the HRS, ELSA, CHARLS, and the Japanese Study of Aging and Retirement, demonstrated that education was an influential socio-economic predictor of healthy ageing.

This highlights the influence of differing national contexts and health systems; for instance, Japan, with its low-cost health services exhibits relatively small socioeconomic inequality in healthy ageing and generally the best healthy ageing profile among the compared nations.^7^ Conversely, the educational inequality in healthy ageing in China was distinctly larger than any other socio-economic inequality, likely reflecting the historical and economic transformations that have deeply stratified educational opportunities and subsequent health outcomes.^7^ The accumulating evidence from these studies paints a clear picture: what happens in early life profoundly shapes health in later life. The potential deficits of healthy ageing, which significantly reduces the wellbeing of older adults and raises healthcare costs, demands a concerted and sustained life course approach to health policy and intervention.

Despite the growing body of evidence, fully leveraging these insights to inform policy and practice, especially across the diverse landscapes of wealthy and developing nations, presents complex challenges including erroneous recall of retrospective information and harmonisation of information from wealthy and developing nations. Further research is needed to refine measurement tools, design targeted and effective interventions and harness the power of longitudinal data for effective health policy.

### Measurement of healthy ageing: healthy ageing index and healthy ageing scale

To deal with the challenges this study stands on the shoulders of two recent multinational studies on healthy ageing scale^6^ and healthy ageing index^7^ and it is closest in purpose with a most recent study by Wu and colleagues which also used the healthy ageing scale.^24^ In comparison, this study differs along two key dimensions: the exposure of childhood poverty and the outcome construction i.e. the healthy ageing scale. Unlike in the most recent study which used parental occupation and education,^24^ in a series of recent multinational studies on the life course shaping of health in later life,^12,14,15,19^ the exposure of early life adversity is a latent construct with multiple items or indicators including housing quality and overcrowding –indicators that are singled out in the WHO Framework to implement a life course approach in practice (:14).^5^

Over the last couple of decades measures of healthy ageing has not settled to a consensus but it has generally been measured in two main approaches. First by measuring multiple organ functions and comparing them with clinical cut-offs, then summing the scores or counting the affirmative comparisons. There may be variations in how comparisons are made but essentially this counting approach relies directly on observed measures. See for example the healthy ageing phenotype and more recently healthy ageing index.^7,8,25^ In contrast, a recent latent approach exemplified in a couple of studies of the ATHLOS project and the Health and Retirement Study relies on latent constructs. For example, the ATHLOS project applied a latent trait model (formulated in 1968)^26^ to 41 observed measurements when deriving the latent trait of healthy ageing. The trait parameter is interpreted as the healthy ageing scale with higher means healthier. It is notable that ATHLOS’ healthy ageing scale does not include mental health while the healthy ageing index does.

To avoid yet another measure of healthy ageing, this study follows closely the latent approach of ATHLOS by applying more recent generalised latent trait model (proposed in 2000)^27^ only this time it includes mental health functions, an important condition in late life.^13,14,28^

Because the study aims not to derive a scale of healthy ageing but to estimate the strength of association between healthy ageing with childhood poverty under a wide variety of national circumstances, I follow the prior practice of focusing on the association. Courtin and colleagues for example estimated the association between mental health and socioeconomic position in older adults over 50 years, comparing and US (HRS) and Europe (SHARE).^29^ Even though HRS and SHARE use different instruments (Center for Epidemiologic Study – Depression and Euro-Depression scales), resulting in important difference in prevalence of depression case, the authors nevertheless proceeded with the estimation of the association between mental health and socioeconomic position. Similarly, Tampubolon and Maharani estimated the association between allostatic load trajectories and their risk factors in US (HRS) and England (ELSA) despite the two longitudinal cohorts collecting different blood biomarkers.^30^ In sum, my focus is not on the difference in averages of healthy ageing scale across nations but on the strengths of its associations with childhood poverty.

### A new healthy ageing scale: from latent trait model to generalised latent trait model

Because the healthy ageing scale has been used in many countries and the latent trait (healthy ageing) has been found to be useful, I start with the healthy ageing scale and proceed with two modifications and call the result healthy ageing scale (modified).^12–17,20,31,32^ First, I include an item on mental health, specifically CESD and Euro-D. The second and more important advance has to do with the latent construction. The healthy ageing scale (original) proceeded by dichotomising *all* continuous items at the first quartile (:883)^6^ in order to make the items fit the latent trait model of Birnbaum (1968).^26^ This practice has long been suspect - it is well known that dichotomising continuous variables leads to loss of information and inconsistent estimation.^33–36^ The table below shows the number of items that are exposed to such risks. This is doubly acute for multinational studies like this (32 nations) because variation in a continuous variable e.g. depression is likely to vary considerably across countries. Already in 1983 Cohen wrote (:249), “since methods are available for making use of all the original scale information there is no reason to sustain them.”^33^ The warning is regular^34,36^ and more recently MacCallum and colleagues wrote (:19), “it is rarely defensible and often will yield misleading results.”^35^ There is another reason to use a more modern *generalised* latent trait model, a direct descendant of Birnbaum’s 1968 latent trait model used in the healthy ageing scale (original) – it is capable of ingesting all scale information not only dichotomous or binary.^27^ The generalised latent trait model has been available since 2000; for details and comparisons see Birnbaum^26^ and Moustaki and Knott.^27^ I use this generalisation to derive a healthy ageing scale (modified) with similar set of items except for the addition of mental health.

In sum, the construction of a modified healthy ageing scale is guided by two principles. First, pragmatism^6,7,37^ so I start with good bases (healthy ageing scale and healthy ageing index).

Second, I apply recent methodological developments.^27,38–43^ The items for constructing the scale are shown in the table below.

### Aims

I aim to explore the associations between childhood poverty and healthy ageing scale under the new WHO Life Course Framework, comparing the experiences of wealthy and developing nations because findings on healthy ageing has been deeply unbalanced across these nations.^4,44^This raises the following questions:

- Does childhood poverty directly associate with healthy ageing in wealthy and developing nations around the world even after parental and contemporaneous risk factors are considered?
- How can ongoing longitudinal cohort studies be used to better capture variations in healthy ageing across diverse global populations?

## MATERIAL AND METHOD

The outcome is healthy ageing scale (modified) which is based on ATHLOS’ healthy ageing scale with two modifications.^6^ First, it includes depression scale (CESD or EuroD) for the psychology domain; second, it replaces Birnbaum’s latent trait model (1968) with Moustaki and Knott’s generalised latent trait model (2000) which relaxes the items’ metric.^26,27^ Unlike the Birnbaum’s model, the generalised model admits all metrics: binary, ordered and continuous of various distributions. Depression scale for instance has a gamma distribution. For each cohort the model is fitted to derive a latent trait which, following the ATHLOS project,^6^ is called the healthy ageing scale (modified) with mean zero and standard deviation (SD) one.

The key exposure is childhood poverty, a latent construct of error-prone childhood information or indicators collected in life history interviews, following ample precedents.^13–17,19,20,45,46^ It is twofold: poor, non-poor. The childhood information or indicators are collected in a table below, again following prior practice.^13–15,19,47^ The other controls include parental socioeconomic or occupational position (twofold: low, high) following prior practice.^14,15,19,20,24,48^ A set of confounders or social determinants for adjustment is included as standard in the empirical literature on ageing including age, sex, education, wealth, marital status and country.^13–15,49,50^ Significance is set at 5% and all estimation is done with Latent Gold syntax^61.51,52^

## RESULTS

### Summary statistics of the analytic sample

The analytic sample comprises 80,319 adults aged 50+ drawn from six well-known longitudinal cohorts (HRS, ELSA, SHARE, TILDA, CHARLS, IFLS), with a balanced age profile (mean 65.6 years, SD 9.5) and a modest female majority (45,476 women; 56.6%). Life course exposure of interest (childhood poverty) is common: nearly one in four participants are classified as childhood poor (24.5%), with a higher prevalence among women (25.2%) than men (23.6%). At baseline, the Healthy Ageing Scale (HAS, modified) shows a small male advantage (men: mean 0.2; women: −0.1), consistent with sex differences observed in functional and mobility domains in many cohorts. Educational attainment is heterogeneous across nations but, in aggregate, 40.4% report up-to-primary education, 37.8% have completed high school, and 21.8% have college or higher. This gradient presents the sizeable education coefficients in multivariable models and mirrors cross-national disparities in access to schooling among the older cohorts under study. Current socioeconomic position (wealth quartiles) remains strongly skewed: 19.7% of the sample are in the bottom wealth quartile, with a larger concentration among women (21.8% vs. 17.0% in men). The intergenerational association is captured by parental socioeconomic/occupational position, where roughly 30% report low parental SEP -consistent with post-war and transitional-economy cohorts. Marital composition varies by sex: men are more often married/partnered (75.7% vs. 60.1%), whereas women are more often separated/widowed (35.4% vs. 18.8%), a familiar pattern reflecting sex differences in longevity. Geographically, Europe supplies the bulk of observations (via SHARE and TILDA), with notable contributions from the USA (8.0% of the sample), China (11.7%) and Indonesia (3.9%). Ireland and England together account for 14.4% via TILDA and ELSA. The spread across 32 nations ensures ample between-country variation in social histories (e.g., welfare regimes, rapid development) and health systems (NHS-type, mixed, private), which is then absorbed in fixed-effects estimation. Taken together, Table 3 indicates (i) substantial heterogeneity in socioeconomic conditions across the life course, (ii) a non-trivial burden of childhood poverty, and (iii) sex, education, and wealth patterns that are plausibly linked to healthy ageing. These features provide statistical power and external validity for modelling associations between childhood poverty and later-life healthy ageing while underscoring the need to adjust for contemporaneous socioeconomic factors and parental socioeconomic position.

**Table 1.**
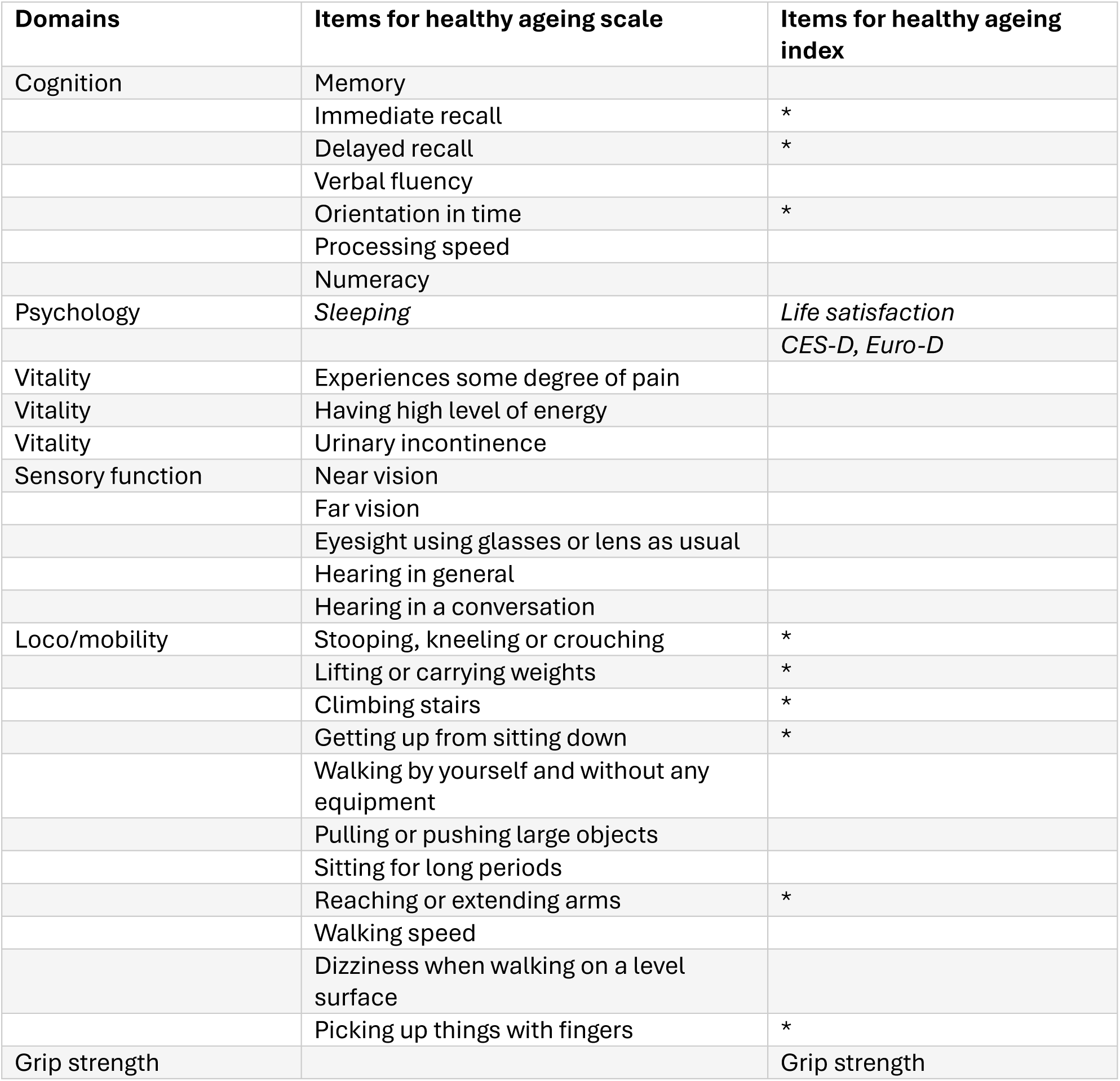
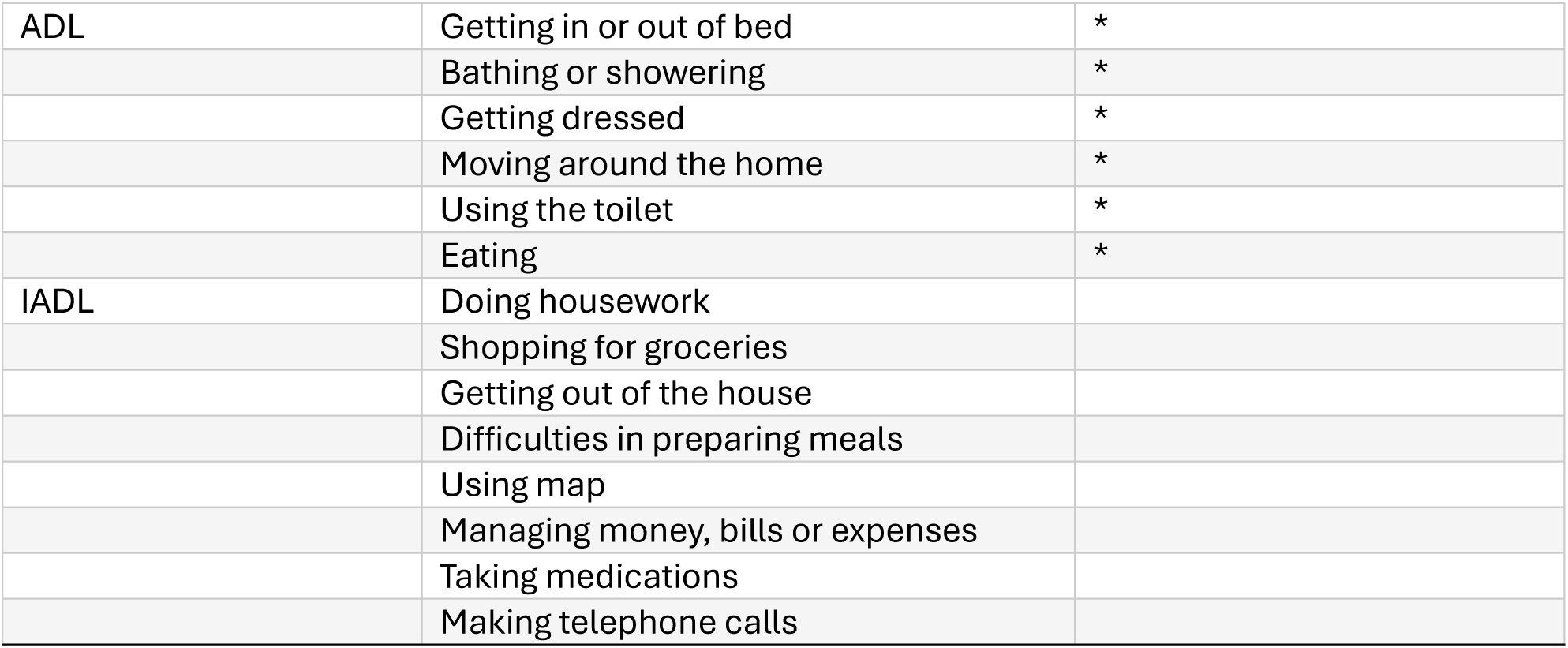
Items for healthy ageing scale^6^ and for healthy ageing index^7^

| Domains | Items for healthy ageing scale | Items for healthy ageing index |
| --- | --- | --- |
| Cognition | Memory |  |
|  | Immediate recall | * |
|  | Delayed recall | * |
|  | Verbal fluency |  |
|  | Orientation in time | * |
|  | Processing speed |  |
|  | Numeracy |  |
| Psychology | *Sleeping* | *Life satisfaction* |
|  |  | *CES-D, Euro-D* |
| Vitality | Experiences some degree of pain |  |
| Vitality | Having high level of energy |  |
| Vitality | Urinary incontinence |  |
| Sensory function | Near vision |  |
|  | Far vision |  |
|  | Eyesight using glasses or lens as usual |  |
|  | Hearing in general |  |
|  | Hearing in a conversation |  |
| Loco/mobility | Stooping, kneeling or crouching | * |
|  | Lifting or carrying weights | * |
|  | Climbing stairs | * |
|  | Getting up from sitting down | * |
|  | Walking by yourself and without any equipment |  |
|  | Pulling or pushing large objects |  |
|  | Sitting for long periods |  |
|  | Reaching or extending arms | * |
|  | Walking speed |  |
|  | Dizziness when walking on a level surface |  |
|  | Picking up things with fingers | * |
| Grip strength |  | Grip strength |
| ADL | Getting in or out of bed | * |
|  | Bathing or showering | * |
|  | Getting dressed | * |
|  | Moving around the home | * |
|  | Using the toilet | * |
|  | Eating | * |
| IADL | Doing housework |  |
|  | Shopping for groceries |  |
|  | Getting out of the house |  |
|  | Difficulties in preparing meals |  |
|  | Using map |  |
|  | Managing money, bills or expenses |  |
|  | Taking medications |  |
|  | Making telephone calls |  |

**Table 2.** Retrospective childhood information in HRS, ELSA, SHARE, TILDA, CHARLS and IFLS.

| Surveys | HRS & TILDA (age <16) | ELSA & SHARE (age 10) |
| --- | --- | --- |
| **Variable** | Number of rooms<br>Number of people<br>Number of books<br>Ever experienced:<br>being financially poor had to move or be helped because of financial difficulty, had to live with grandparents, father unemployed | Number of rooms<br>Number of people<br>Number of books<br>Physical facilities:<br>indoor toilet, fixed bath,<br>central heating, hot & cold running water |
| Surveys | CHARLS (age <17) | IFLS (age 12) |
| **Variable** | Family member starved to death | Number of rooms |
|  | Had to move due of financial difficulty | Number of people |
|  | Experienced financial hardship | Had electricity |
|  | Living in commune that is unsafe | Had cold running water |
|  | ... less willing to help others | Had indoor toilet |
|  | ... not close knit | Experienced hunger or famine |
|  | ... not clean |  |

**Table 3.** Summary statistics of analytic sample for explaining healthy ageing scale drawn from six longitudinal ageing cohorts (HRS, ELSA, SHARE, TILDA, CHARLS, IFLS).

|  | Male | Female | Total |
| --- | --- | --- | --- |
|  | N=34843 | N=45476 | N=80319 |
| Childhood poor |  |  |  |
| Childhood non-poor | 26625 (76.4%) | 34027 (74.8%) | 60652 (75.5%) |
| Childhood poor | 8218 (23.6%) | 11449 (25.2%) | 19667 (24.5%) |
| Healthy Ageing scale | 0.2 (1.0) | -0.1 (1.0) | -0.0 (1.0) |
| Age | 65.7 (9.2) | 65.5 (9.7) | 65.6 (9.5) |
| Education |  |  |  |
| Up to Primary | 12574 (36.1%) | 19852 (43.7%) | 32426 (40.4%) |
| High school | 14059 (40.3%) | 16292 (35.8%) | 30351 (37.8%) |
| College/more | 8210 (23.6%) | 9332 (20.5%) | 17542 (21.8%) |
| Marital status |  |  |  |
| Single | 1902 (5.5%) | 2056 (4.5%) | 3958 (4.9%) |
| Married/Partnered | 26385 (75.7%) | 27315 (60.1%) | 53700 (66.9%) |
| Sep/Widowed | 6556 (18.8%) | 16105 (35.4%) | 22661 (28.2%) |
| Current wealth |  |  |  |
| Above bottom quartile | 28935 (83.0%) | 35568 (78.2%) | 64503 (80.3%) |
| Bottom quartile | 5908 (17.0%) | 9908 (21.8%) | 15816 (19.7%) |
| Parent socioeconomic position |  |  |  |
| SEP high | 24121 (69.2%) | 31805 (69.9%) | 55926 (69.6%) |
| SEP low | 10722 (30.8%) | 13671 (30.1%) | 24393 (30.4%) |
| Country |  |  |  |
| Ireland | 1965 (5.6%) | 2351 (5.2%) | 4316 (5.4%) |
| England | 3207 (9.2%) | 4019 (8.8%) | 7226 (9.0%) |
| Austria | 742 (2.1%) | 1128 (2.5%) | 1870 (2.3%) |
| Germany | 1660 (4.8%) | 1864 (4.1%) | 3524 (4.4%) |
| Sweden | 1245 (3.6%) | 1459 (3.2%) | 2704 (3.4%) |
| Netherlands | 644 (1.8%) | 842 (1.9%) | 1486 (1.9%) |
| Spain | 1254 (3.6%) | 1547 (3.4%) | 2801 (3.5%) |
| Italy | 1415 (4.1%) | 1737 (3.8%) | 3152 (3.9%) |
| France | 1351 (3.9%) | 1843 (4.1%) | 3194 (4.0%) |
| Denmark | 1294 (3.7%) | 1499 (3.3%) | 2793 (3.5%) |
| Greece | 332 (1.0%) | 431 (0.9%) | 763 (0.9%) |
| Switzerland | 1024 (2.9%) | 1244 (2.7%) | 2268 (2.8%) |
| Belgium | 1499 (4.3%) | 1827 (4.0%) | 3326 (4.1%) |
| Israel | 333 (1.0%) | 486 (1.1%) | 819 (1.0%) |
| Czechia | 1216 (3.5%) | 1891 (4.2%) | 3107 (3.9%) |
| Poland | 1253 (3.6%) | 1578 (3.5%) | 2831 (3.5%) |
| Luxembourg | 377 (1.1%) | 461 (1.0%) | 838 (1.0%) |
| Hungary | 262 (0.8%) | 413 (0.9%) | 675 (0.8%) |
| Slovenia | 937 (2.7%) | 1356 (3.0%) | 2293 (2.9%) |
| Estonia | 1031 (3.0%) | 1825 (4.0%) | 2856 (3.6%) |
| Croatia | 493 (1.4%) | 635 (1.4%) | 1128 (1.4%) |
| Lithuania | 475 (1.4%) | 861 (1.9%) | 1336 (1.7%) |
| Bulgaria | 348 (1.0%) | 520 (1.1%) | 868 (1.1%) |
| Cyprus | 182 (0.5%) | 311 (0.7%) | 493 (0.6%) |
| Finland | 520 (1.5%) | 592 (1.3%) | 1112 (1.4%) |
| Latvia | 267 (0.8%) | 470 (1.0%) | 737 (0.9%) |
| Malta | 335 (1.0%) | 425 (0.9%) | 760 (0.9%) |
| Romania | 513 (1.5%) | 694 (1.5%) | 1207 (1.5%) |
| Slovakia | 431 (1.2%) | 508 (1.1%) | 939 (1.2%) |
| USA | 2527 (7.3%) | 3859 (8.5%) | 6386 (8.0%) |
| Indonesia | 1234 (3.5%) | 1863 (4.1%) | 3097 (3.9%) |
| China | 4477 (12.8%) | 4937 (10.9%) | 9414 (11.7%) |

### Model results for the Healthy Ageing Scale (table 4)

Both fixed-effects and random-effects specifications return nearly identical coefficients, indicating that the main inferences are robust to alternative assumptions about cross-country heterogeneity. The focal coefficient for childhood poverty is −0.140 (z −17.6), implying that, net of covariates, older adults who grew up poor have healthy ageing scores lower by 0.14 SD relative to those who were not poor. In standardised terms, this is a meaningful gap: it is roughly half the magnitude of the female–male difference and a quarter of the college advantage. Sex differences are pronounced: women score −0.276 SD lower than men, all else equal. Some of this may reflect differential exposure to caregiving for instance; the models control for marital status and wealth, strongly suggesting that additional pathways (e.g. unpaid care histories) may be at play. Age has the expected negative slope (−0.0376 per year): over a decade, expected HAS declines by 0.38 SD, consistent with physiological ageing and accumulation of impairments. Education exhibits strong, graded associations: high school completion is associated with +0.294 SD and college or more with +0.534 SD relative to up-to-primary. The college coefficient is large enough to offset the childhood-poverty penalty nearly four times over, highlighting education as a potent lever of resilience. Current material conditions matter as well: being in the bottom wealth quartile associates with −0.255 SD, comparable to nearly two childhood-poverty penalties. Parental background retains an independent, albeit smaller, imprint: low parental occupational position associates with −0.050 SD after controlling for the individual’s own education and wealth, supporting an intergenerational perspective in healthy ageing. In the presence of all these associations, childhood poverty remains significant, suggesting the life course shaping of childhood poverty works beyond the pathway of socioeconomic positions past childhood.

**Table 4.**
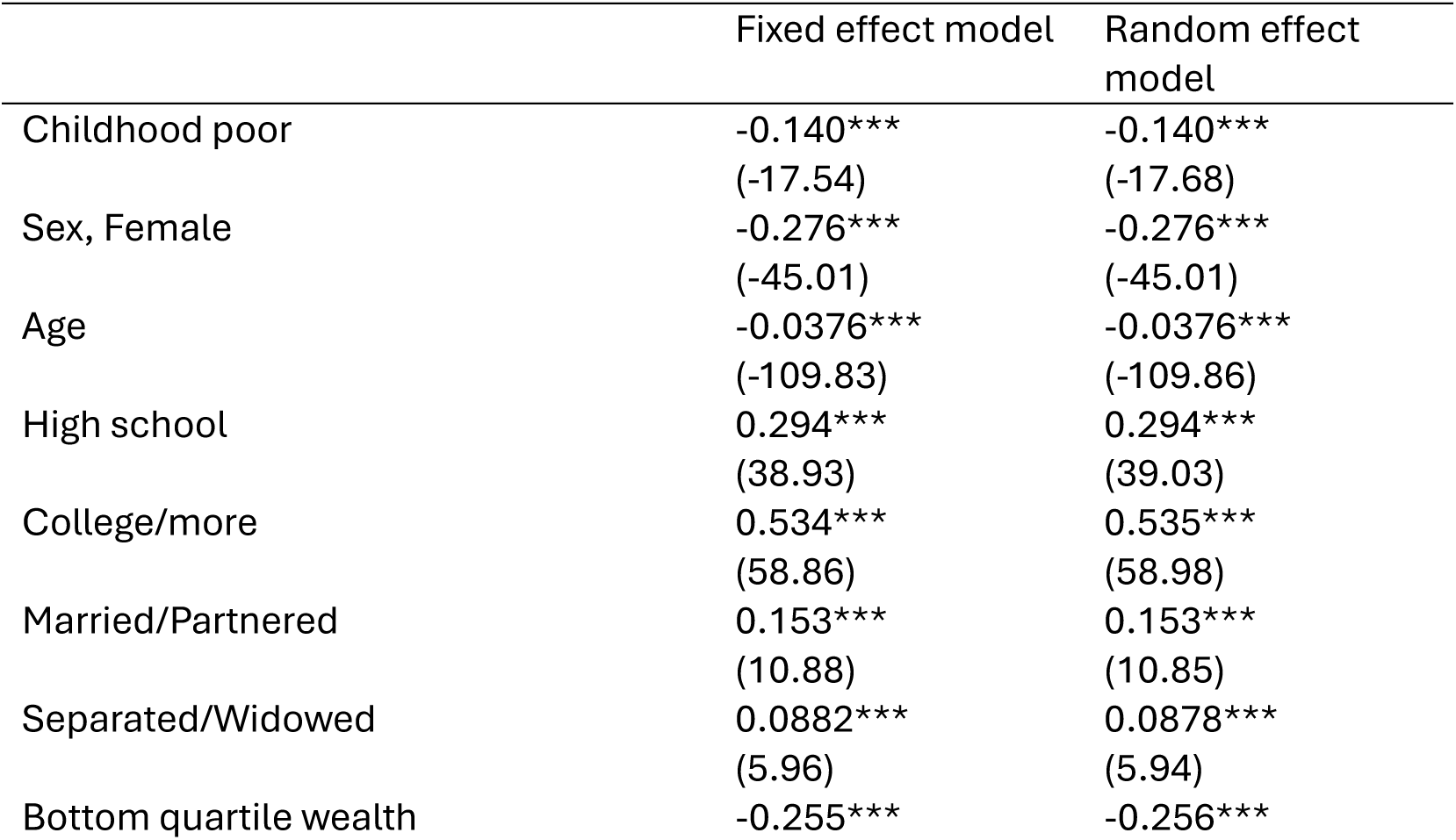
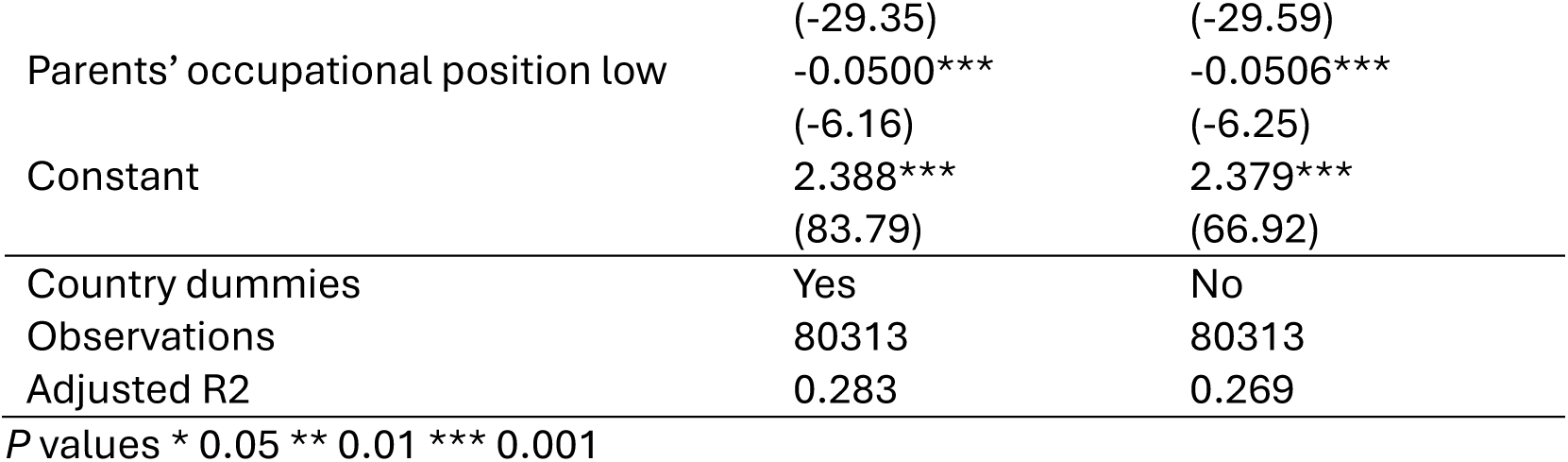
Results of model estimation for healthy ageing scale: fixed effect model and random effect model

Marital status shows positive associations for married/partnered (+0.153 SD) and, interestingly, for separated/widowed (+0.088 SD), which could reflect selective mortality; country controls attenuate unobserved heterogeneity. The fixed-effects model includes country dummies and explains a sizable share of variance (adjusted R²=0.283) for a composite, multidimensional outcome. The near identical fixed- and random-effects estimates, alongside very large z-statistics, underpins the stability of results across modelling choices and strengthens the conclusion that childhood poverty exerts an independent and consistent drag on healthy ageing across the world.

### Marginal plot for ages 70-90

To ease comprehension, I draw marginal predictions for ages 70–90, plotted for each country, showing that in all setting the childhood-poor age profiles (dashed lines) lie below the non-poor profile (solid lines) throughout. This trellis plot shows at a glance a pervasive childhood poverty penalty on healthy ageing. Moreover, both baseline levels (age 70) and slopes vary widely across countries. In some nations the gap is approximately parallel across age (stable disadvantage); elsewhere it widens (cumulative disadvantage). Occasional convergence at extreme ages likely reflects selective mortality or survival bias.

**Figure 1.**
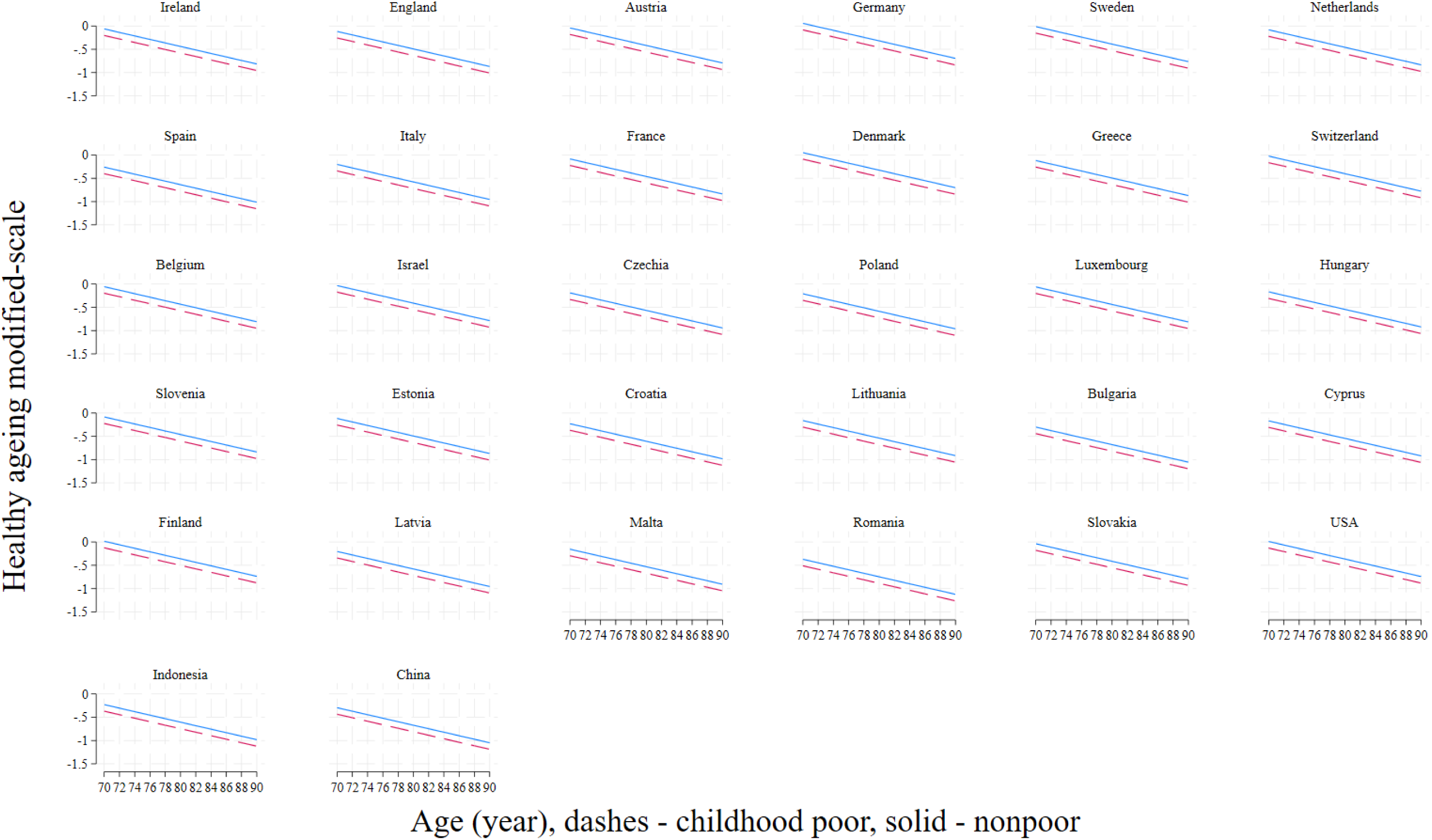
Marginal predictions of Healthy Ageing Scale (modified) for ages 70–90 across 32 nations in six cohorts. Dashed = childhood poor; Solid = non-poor.

### Supplementary Sensitivity Analyses: sex stratification, cohort-specific results and sample non-response patterns

I assess the main results above against three alternatives, first whether the results deviate considerably across the sexes, in each specific cohort and what sample construction potentially affects. In other words, because not all participants agreed to give life history interviews, nor give all responses to healthy ageing items (table 1) the constructed sample (made up of those who agreed to both) maybe biased, say they tended to be younger – the non-response patterns need to be examined (table 7). In table 5, sex-stratified models confirm robustness: childhood poverty lowers healthy ageing for both women (–0.150 SD) and men (– 0.125 SD), with nearly identical age slopes (–0.038 per year), supporting a consistent life course penalty across sexes. Education remains protective (high school +0.298 women; +0.275 men; college +0.53 both), while bottom-quartile wealth is detrimental (women –0.264; men –0.249). Marital status benefits are larger for men (married/partnered +0.208) than women (+0.106), and parental low occupational position has small negative effects in both sexes (–0.05).

**Table 5.** Results of model estimation for healthy ageing scale by sex

|  | Female | Male |
| --- | --- | --- |
| Childhood poor | -0.150***<br>(-14.45) | -0.125***<br>(-10.17) |
| Age | -0.0377*** | -0.0378*** |
|  | (-83.54) | (-71.14) |
| High school | 0.298*** | 0.275*** |
|  | (29.68) | (23.87) |
| College/more | 0.535*** | 0.525*** |
|  | (43.69) | (38.48) |
| Married/Partnered | 0.106*** | 0.208*** |
|  | (5.46) | (10.10) |
| Separated/Widowed | 0.0388** | 0.142*** |
|  | (1.97) | (6.21) |
| Wealth: bottom quartile | -0.264*** | -0.249*** |
|  | (-23.79) | (-17.87) |
| Parental occupational position low | -0.0524*** | -0.0479*** |
|  | (-4.94) | (-3.81) |
| Constant | 2.188*** | 2.321*** |
|  | (57.45) | (55.21) |
| Observations | 45472 | 34841 |
| Adjusted $R^2$ | 0.291 | 0.234 |

In table 6 set, cohort-specific estimates reaffirm the main result but also reveal meaningful heterogeneity. The childhood-poverty penalty is strongest in China/CHARLS (–0.311), sizeable in SHARE (–0.168), IFLS (–0.187), and HRS (–0.184), moderate in TILDA (–0.133), and smallest in ELSA (–0.063). Female disadvantage is largest in CHARLS (–0.465), while the wealth penalty peaks in HRS (–0.489) and ELSA (–0.406) but is attenuated in IFLS (–0.094) and CHARLS (–0.148). Education returns are strongest in IFLS and CHARLS. Overall, findings are robust, with variations plausibly linked to welfare regimes, history and social development.

**Table 6.a.** Results of model estimation for healthy ageing scale per cohort – Europe (SHARE)

|  | Europe (SHARE) |
| --- | --- |
| Childhood poor | -0.168*** |
|  | (-18.43) |
| Sex, Female | -0.244*** |
|  | (-31.22) |
| Age | -0.0419*** |
|  | (-97.20) |
| High school | 0.291*** |
|  | (32.94) |
| College/more | 0.550*** |
|  | (51.95) |
| Married/Partnered | 0.132*** |
|  | (7.54) |
| Separated/Widowed | 0.0968*** |
|  | (5.15) |
| Wealth: bottom quartile | -0.285*** |
|  | (-28.10) |
| Parental occupational position low | -0.0510*** |
|  | (-5.72) |
| Constant | 2.679*** |
|  | (78.10) |
| Observations | 49880 |
| Adjusted $R^2$ | 0.302 |

*t* statistics in parentheses
\* $p < 0.1$ , \*\* $p < 0.05$ , \*\*\* $p < 0.01$

**Table 6.b.**
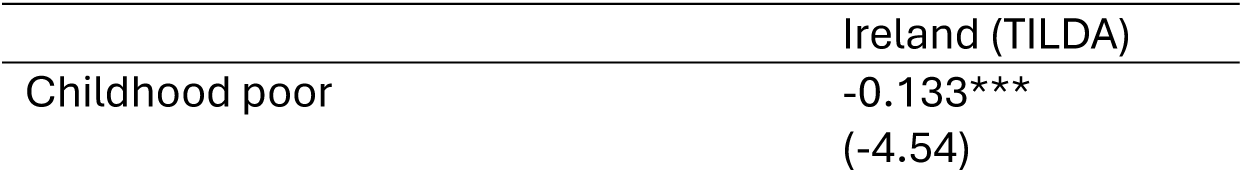
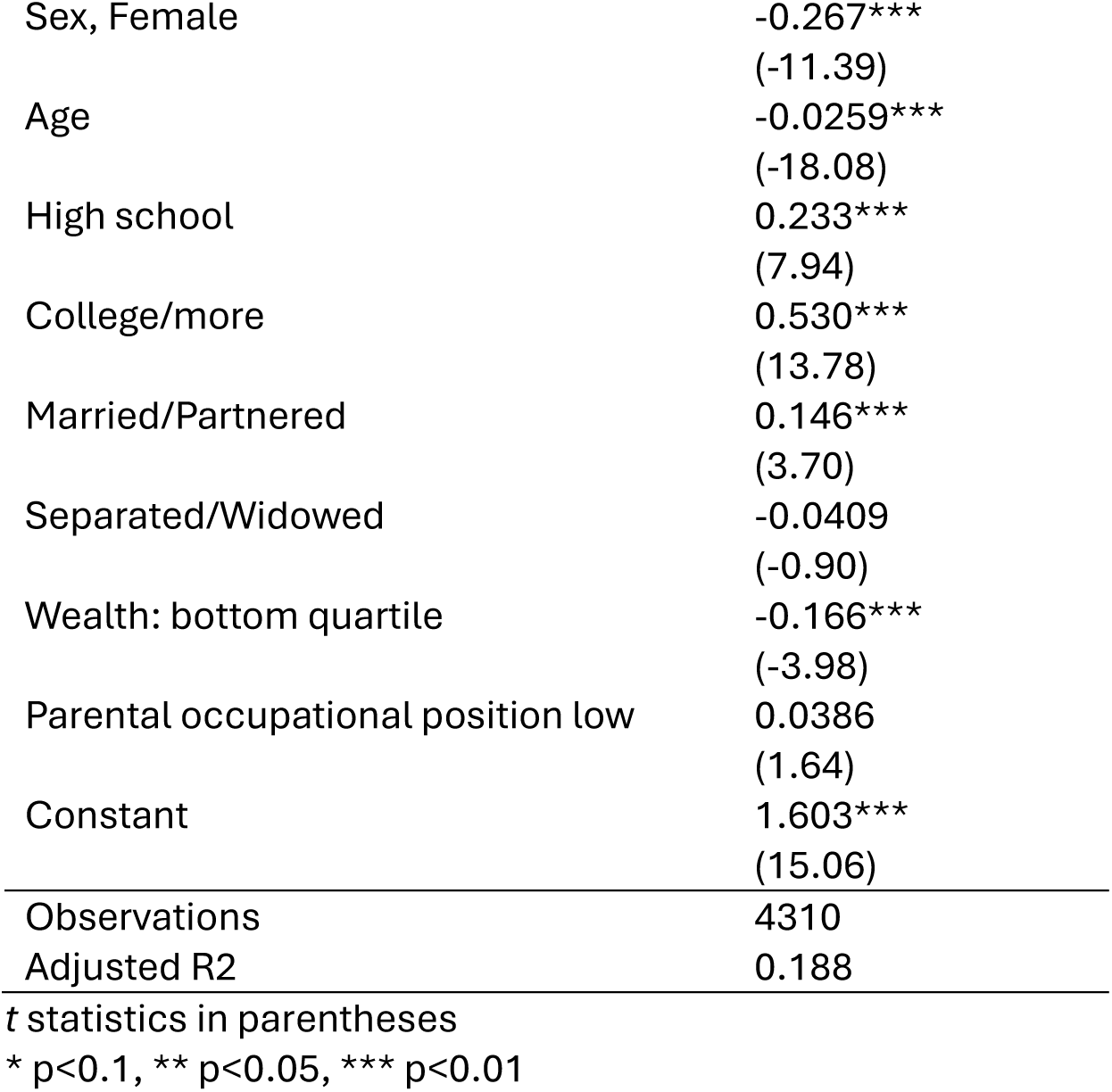
Results of model estimation for healthy ageing scale per cohort – Ireland (TILDA)

|  | Ireland (TILDA) |
| --- | --- |
| Childhood poor | -0.133*** |
|  | (-4.54) |
| Sex, Female | -0.267***<br>(-11.39) |
| Age | -0.0259***<br>(-18.08) |
| High school | 0.233***<br>(7.94) |
| College/more | 0.530***<br>(13.78) |
| Married/Partnered | 0.146***<br>(3.70) |
| Separated/Widowed | -0.0409<br>(-0.90) |
| Wealth: bottom quartile | -0.166***<br>(-3.98) |
| Parental occupational position low | 0.0386<br>(1.64) |
| Constant | 1.603***<br>(15.06) |
| Observations | 4310 |
| Adjusted R2 | 0.188 |

*t* statistics in parentheses
\* $p < 0.1$ , \*\* $p < 0.05$ , \*\*\* $p < 0.01$

**Table 6.c.** Results of model estimation for healthy ageing scale per cohort – England (ELSA)

|  | England (ELSA) |
| --- | --- |
| Childhood poor | -0.0627***<br>(-2.61) |
| Sex, Female | -0.225***<br>(-11.32) |
| Age | -0.0305***<br>(-28.48) |
| High school | 0.238***<br>(9.55) |
| College/more | 0.425***<br>(16.86) |
| Married/Partnered | 0.108**<br>(2.56) |
| Separated/Widowed | -0.0161<br>(-0.36) |
| Wealth: bottom quartile | -0.406***<br>(-16.84) |
| Parental occupational position low | -0.159***<br>(-3.63) |
| Constant | 2.019***<br>(23.26) |
| Observations | 7226 |
| Adjusted R2 | 0.261 |

*t* statistics in parentheses
\* $p < 0.1$ , \*\* $p < 0.05$ , \*\*\* $p < 0.01$

**Table 6.d.** Results of model estimation for healthy ageing scale per cohort – US (HRS)

|  | USA (HRS) |
| --- | --- |
| Childhood poor | -0.184***<br>(-5.32) |
| Sex, Female | -0.251***<br>(-10.77) |
| Age | -0.0254***<br>(-22.39) |
| High school | 0.0597<br>(1.52) |
| College/more | 0.366***<br>(9.52) |
| Married/Partnered | 0.145<br>(1.20) |
| Separated/Widowed | 0.0473<br>(1.12) |
| Wealth: bottom quartile | -0.489***<br>(-14.40) |
| Parental occupational position low | NA<br>NA |
| Constant | 1.731***<br>(18.42) |
| Observations | 6386 |
| Adjusted R2 | 0.158 |

*t* statistics in parentheses
\* $p < 0.1$ , \*\* $p < 0.05$ , \*\*\* $p < 0.01$
99% of parental occupation is not released for public use.

**Table 6.e.**
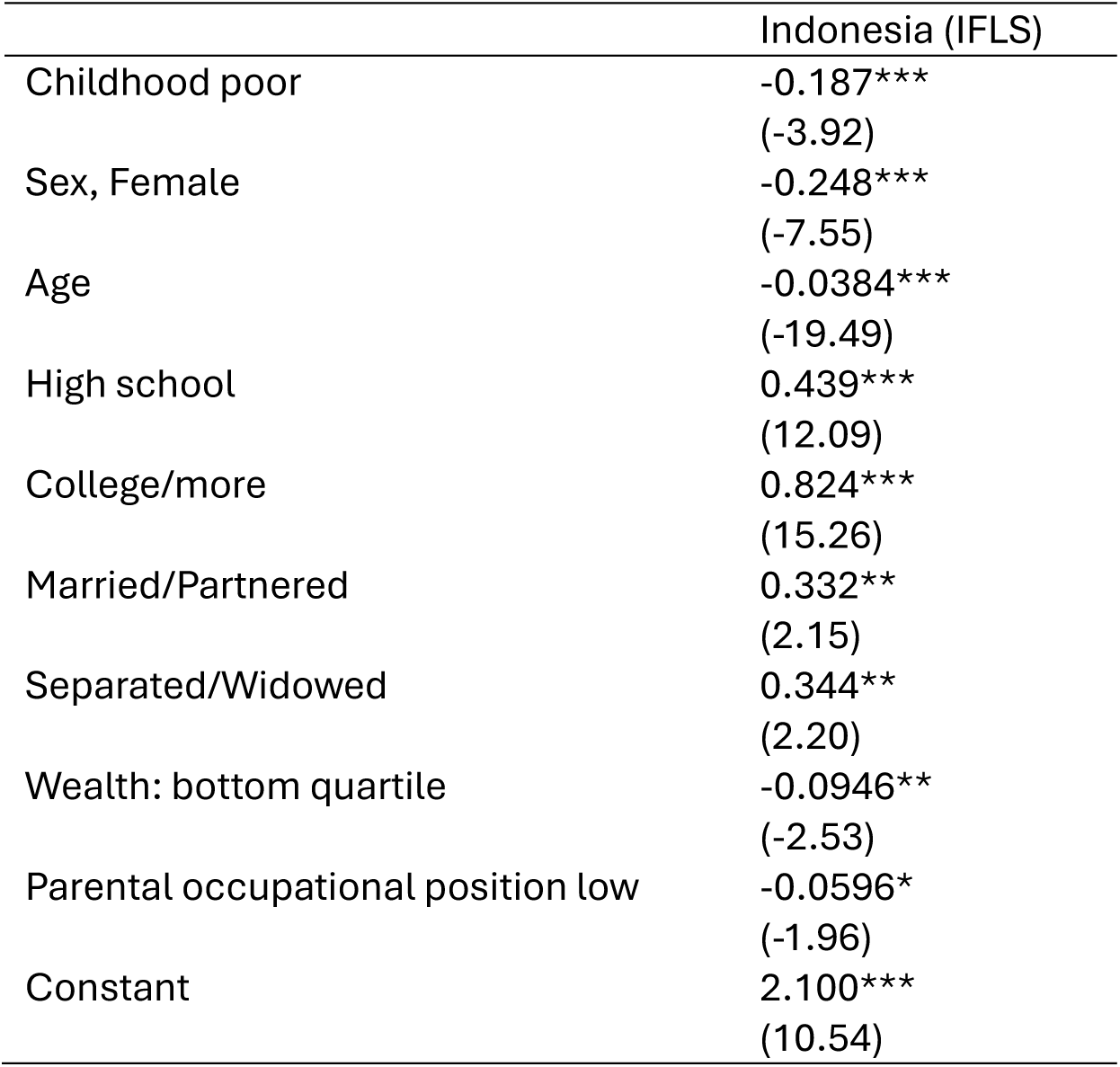
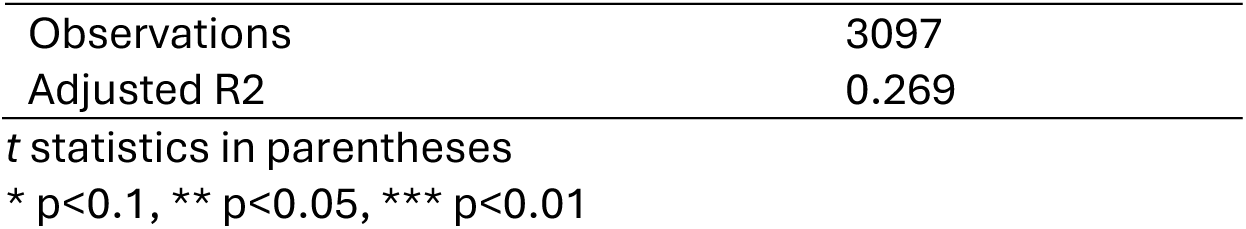
Results of model estimation for healthy ageing scale per cohort – Indonesia (IFLS)

**Table 6.f.**
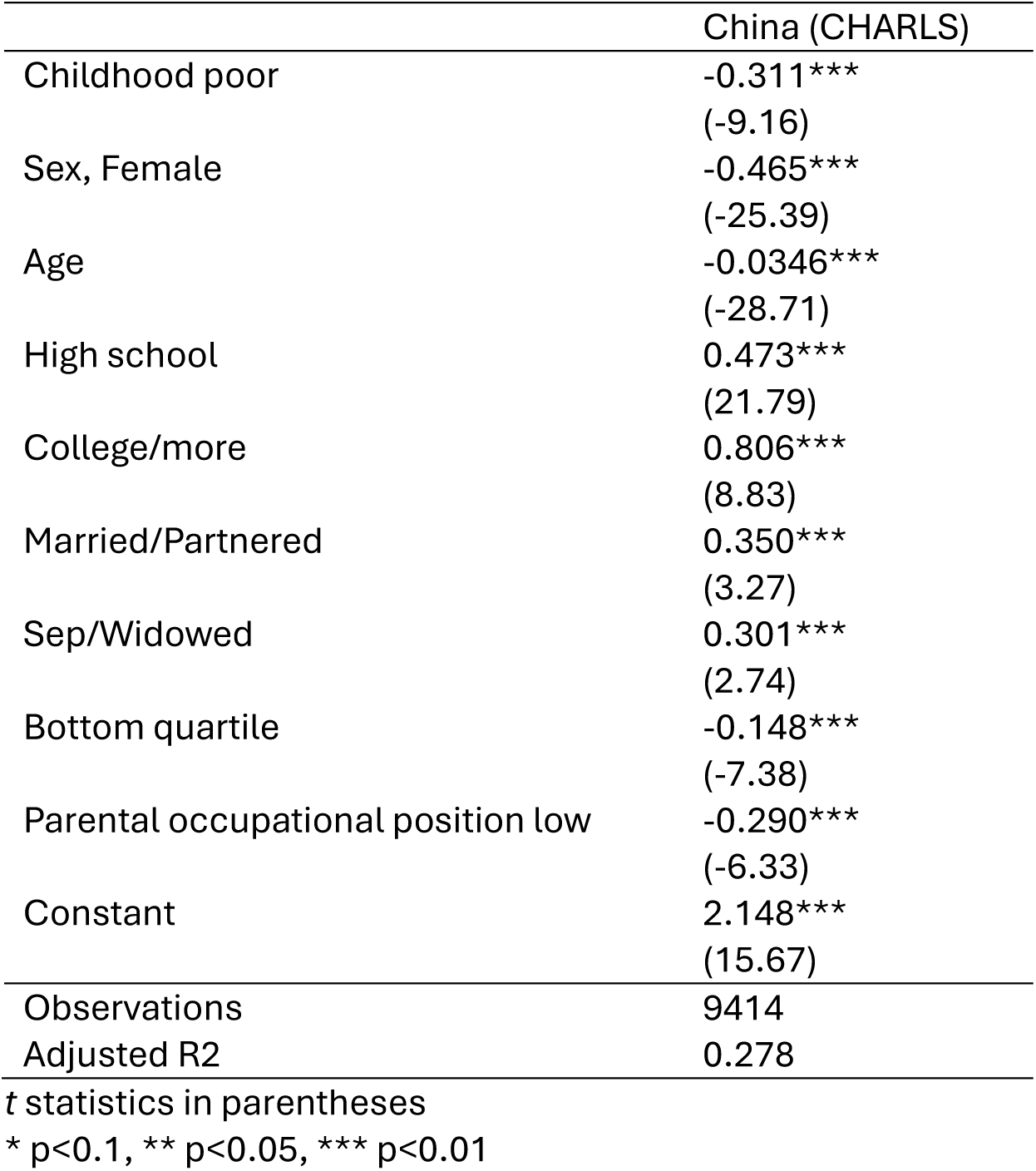
Results of model estimation for healthy ageing scale per cohort – China (CHARLS)

|  | China (CHARLS) |
| --- | --- |
| Childhood poor | -0.311***<br>(-9.16) |
| Sex, Female | -0.465***<br>(-25.39) |
| Age | -0.0346***<br>(-28.71) |
| High school | 0.473***<br>(21.79) |
| College/more | 0.806***<br>(8.83) |
| Married/Partnered | 0.350***<br>(3.27) |
| Sep/Widowed | 0.301***<br>(2.74) |
| Bottom quartile | -0.148***<br>(-7.38) |
| Parental occupational position low | -0.290***<br>(-6.33) |
| Constant | 2.148***<br>(15.67) |
| Observations | 9414 |
| Adjusted R2 | 0.278 |

*t* statistics in parentheses \* $p < 0.1$ , \*\* $p < 0.05$ , \*\*\* $p < 0.01$

**Table 7.** Non-response patterns across analytic vs declined sample in HRS, ELSA, SHARE, TILDA, CHARLS & IFLS.

| Survey | Variable | Declined sample | Analytic sample |
| --- | --- | --- | --- |
| **HRS** | Female | 60% | 59% |
|  | Male | 40% | 41% |
| | | $\chi^2_1 = 1.0$ | $p = 0.30$ |
| | Mean age | 65.1 year | 68.4 year<br>$p < 0.001$ |
| **ELSA** | Female | 74% | 55% |
|  | Male | 26% | 45% |
| | | $\chi^2_1 = 87.8$ | $p < 0.001$ |
| | Mean age | 65.3 year | 54.1 year<br>$p < 0.001$ |
| **SHARE** | Female | 56% | 58% |
|  | Male | 44% | 42% |
| | | $\chi^2_1 = 8.9$ | $p = 0.003$ |
|  | Mean age | 67.3 year | 68.5 year |
| | | | $p < 0.001$ |
| **CHARLS** | Female | 48% | 53% |
|  | Male | 52% | 47% |
| | | $\chi^2_1 = 36.8$ | $p < 0.001$ |
|  | Mean age | 62.2 year | 64.1 year |
| | | | $p < 0.001$ |
| **IFLS** | Female | 59% | 53% |
|  | Male | 41% | 47% |
| | | $\chi^2_1 = 19.0$ | $p < 0.001$ |
|  | Mean age | 70.7 year | 59.8 year |
| | | | $p < 0.001$ |
| **TILDA** | Female | 89% | 54% |
|  | Male | 11% | 46% |
| | | $\chi^2_1 = 155$ | $p < 0.001$ |
|  | Mean age | 49 year | 63.5 year |
| | | | $p < 0.001$ |

#### Non-response patterns for retrospective interviews

The analytic sample comprises prospective participants who agreed to retrospective life history interviews. Some declined and there may be significant differences between the analytic *vs* declined sample, tested using 𝜒^2^and *t* tests for nominal and continuous variables as appropriate (table 7). In the HRS the analytic sample has 59% females and 41% males, with 𝜒^2^ = 1.0 and 𝑝 value = 0.30; a pattern also observed in the sample who declined the interviews. In contrast, in other cohorts the analytic samples females are more likely to agree to the interviews. Meanwhile in the HRS the analytic sample has older participants, and this difference is significant while in ELSA the opposite is true while also significant. In sum, there are no strong patterns of participation in retrospective interviews around the world.

## DISCUSSION

The childhood poverty penalty provides a clear, positive-orientation analogue to prior deficit-oriented outcomes: where earlier studies observed higher depression, greater probable sarcopenia, higher frailty or higher multimorbidity among those raised in poverty,^13–15,19^ we now observe less healthy ageing. In standard-deviation unit, the penalty is roughly half the sex gap and one-quarter of the college advantage. The robustness of this association to model specification and to sex-stratification underscores a widely shared life course imprint of material deprivation despite contemporary socioeconomic attainment e.g. education and wealth. This is consistent with epigenetic evidence I supplied elsewhere showing the childhood poor aged epigenetically faster.^14^

Cross-country heterogeneity is informative rather than contradictory. Higher baselines and gentler declines -visible in several northern European settings- are consistent with comprehensive primary care, rehabilitation and social care supports, while steeper slopes in other settings may reflect unmet need for chronic-disease management, limited rehabilitation coverage or less friendly built environments for ageing. The cohort-by-cohort estimates further suggest historically rooted differences: e.g., the strongest childhood poverty penalty in China plausibly aligns with cohort exposure to the Great Leap Forward and rapid social transition,^53–55^ whereas smaller penalties in England may reflect longer-standing welfare protections.^14,15,19,56^

### Advancing a faithful implementation of the WHO life course framework

This study provides one of the clearest presentations of the WHO life course framework for healthy ageing—treating health in later life as the product of cumulative exposures, critical periods and interacting social–biological pathways across the entire span from childhood to old age, while measuring the outcome as functional ability rather than mere disease absence.^5^ I first constructed a modified healthy ageing scale as a latent trait that integrates cognition, locomotion, sensory function, vitality, activities of daily living and mental health; then modelled childhood poverty as a latent construct to capture material disadvantages highlighted in the WHO framework (page 14; housing quality, overcrowding, utilities, financial hardship);^5^ further, examined associations after adjusting for parental socioeconomic position, age, sex, education, wealth, marital status and country dummies, thereby locating later-life functioning within a web of structural determinants rather than individual risk factors alone. In doing so, the study extends recent cross-country life course evidence on single domains such as episodic memory or grip strength to a positive, composite outcome in which higher scores mean healthier ageing, fully aligned with WHO’s emphasis on functionings.^2,5^ The core result -a childhood poverty penalty on healthy ageing- provides a coherent bridge from deficit-oriented outcomes to a capabilities-oriented metric of later-life functioning.

### Breadth and generalising beyond 32 nations

A second advance is geographic reach. By pooling six longitudinal cohorts -HRS (USA), ELSA (England), SHARE (27 European countries), TILDA (Ireland), CHARLS (China), IFLS (Indonesia)-I study the life course imprint of childhood poverty across 32 nations that differ widely in income levels, welfare regimes, epidemiological transitions and health systems.^56^ The pooled models, country fixed effects, and the trellis of marginal predictions for ages 70–90 jointly show that the childhood-poverty penalty is ubiquitous: predicted HAS for the childhood poor (dashed) lies below the non-poor (solid) at the same age. Yet the levels (baselines at 70) and slopes (declines to 90) vary substantially, reflecting differences in prevention, rehabilitation, social protection, cohort shocks and diagnostic capacity, exactly the heterogeneity the WHO framework expects. This combination -consistent penalty, heterogeneous trajectories- enhances external validity while cautioning against one-size-fits-all prescriptions.

At the same time, I document meaningful cross-cohort variation in the magnitude of the penalty: strongest in China, moderate in Ireland and smallest in England. These differences plausibly reflect cohort-specific histories (e.g., Great Leap Forward legacy in China),^20,53,55^ social policy, and measurement contrast (e.g., wealth reporting, depression coverage) even after harmonisation and fixed-effects controls. Such heterogeneity echoes earlier cross-country patterns reported for frailty, multimorbidity and cognitive functions and probable sarcopenia,^14,15,19^ but now in healthy ageing scale (modified), which captures the net functional consequences of those domain-specific deficits.

### Addressing the imbalance in the evidence base

A recurring limitation in the literature is that life course evidence is over-represented in high-income Western settings; consequently, global strategies risk being extrapolated from contexts with dense data and strong health systems to those with sparse data and constrained systems.^44^ This study directly reduces that imbalance by adding China and Indonesia to a trans-Atlantic nations and by reporting pooled as well as cohort-by-cohort estimates. For example, educational gradients are steepest where schooling expanded later and faster (China and Indonesia), consistent with structural change in human capital formation; wealth penalties are large where financial protection is less universal (US), illustrating that current socio-economic conditions continue to shape functionings in later life. By placing these results alongside Europe- and US-centric findings, the study broadens the empirical canvas on which the WHO Decade of Healthy Ageing^3^ can paint.

### Recall error: from problem recognition to methodological solution

Unlike most studies of childhood conditions and their association with older adults’ health, this study is deliberately self-critical about retrospective childhood measures. It quantifies recall error using independent validations: in England, only one-in-three of 50- or 62-year-olds correctly recalled both number of rooms and people in their childhood versus records collected from their mothers decades earlier. It is unsafe to treat childhood recollections as error-free.

Consequently, following recent practices childhood poverty is a latent construct using diverse indicators (overcrowding, facilities, financial hardship, famine/starvation). This practice is here brought to bear on healthy ageing and across a wider set of countries, improving inference without discarding valuable but imperfect data.

Beyond bias control, the latent construct approach enables comparability when indicator sets differ (e.g., ELSA/SHARE ask about running hot water and central heating; HRS emphasises financial events; CHARLS includes death from starvation). By targeting the association while controlling for country effects, I can compare strength and direction of life course links even if prevalence is not directly harmonised -the argument used in cross-national studies of depression and allostatic load.^28,29^ The novel step here is to apply that argument to a modified HAS, preserving metric information via a generalised latent trait model rather than dichotomising continuous items, thereby aligning statistical practice with the conceptual complexity of healthy ageing.

### From “long arm” mechanisms to “healthy ageing” consequences

I have proposed and supported biological pathways -notably epigenetic aging measured by methylation clocks- to explain how childhood poverty “gets under the skin”.^14^ Here I extend the consequences of those pathways to a composite functional outcome: the same early-life hardship that predicted higher depression, worse muscle function, frailty or multimorbidity now predicts lower healthy ageing decades later, even after accounting for parental and older adults’ socioeconomic positions. This coherence across domains strengthens the causal plausibility of life course mechanisms while remaining cautious about causal claims. The convergence of epidemiologic (childhood poverty penalty), socio-economic (education/wealth gradients) and biological (epigenetic age) signals is noteworthy: a single, conceptually unified story can now be told about how childhood conditions become embedded and what that embedding looks like when the outcome is capability in later life.

### Strengths and limitations

This work has real limitations and some strengths. This study is subject to several limitations that warrant careful consideration. First, the analytical framework employed herein is relatively simple and does not constitute a full structural model.^17^ While precedents for such simplified approaches exist,^13,14,57^ the structural alternative has led some scholars to posit that the inclusion of additional covariates might attenuate the observed influence of childhood conditions. This proposition remains an empirical question. Nonetheless, the imperative persists: a more comprehensive framework should be pursued when harmonized data on youth and adult conditions across wealthy and developing nations become available. Second, the study’s focus on material deprivation in childhood excludes psychological dimensions, such as parent-child bonding relationships. These aspects are central to child development and represent a distinct stage within the broader life course framework. Their omission constitutes a notable limitation. Third, the inclusion of country fixed effects serves to control for unobserved heterogeneity across national contexts, including historical trajectories and health systems.

Although this approach follows precedent,^13,14,17^ it may be deemed insufficient. Accordingly, country-specific analyses for the 32 nations in this study are deferred to a future monograph.

Fourth, the observational nature of the study design precludes causal inference.

Elsewhere I noted that the absence of random assignment to childhood material conditions introduces potential confounding, as individuals who experienced poverty in childhood may differ systematically from those who did not.^14,15^ Moreover, survivor bias may be present, as some individuals exposed to childhood poverty may not have lived to participate in ageing surveys. While this bias may suggest a directional effect, its magnitude remains unknown.

Furthermore, the feasibility of randomised designs in this context is questionable, given the extended temporal gap -often spanning half a century- between exposure and outcome. Such a duration increases the likelihood of behavioural adaptation following randomisation, thereby undermining causal inference. Despite these constraints, support for the life course framework may be bolstered through cross-national design.

The strengths of this study merit emphasis. First, faithful WHO life course implementation. The analysis aligns measurement (modified HAS), key exposure (latent childhood poverty with indicators of overcrowding and housing qualities), and covariate structure (parental occupational position, age, sex, education, wealth, marital status, country fixed effects) with the WHO framework’s emphasis on functional ability and social determinants, avoiding a disease-only lens. Second, an exceptional geographic scope for the key exposure. Six cohorts across 32 nations provide both breadth and heterogeneity, giving confidence to generalisation and revealing country variation (levels, slopes) in later-life functioning. Third, methodological discipline on recall error. There is scant evidence on recall error in retrospective information and when evidence is found at age 50 and beyond it is sizeable - two-thirds got childhood conditions wrong. This is addressed with latent constructs and corrected standard errors, improving validity without discarding data. Fourth, metric-preserving outcome construction. Using a *generalised* latent trait model retains information and suits multinational analyses where item distributions differ, aligning methodology with conceptual nuance. Last, robustness. Fixed vs random effects yield identical coefficients; sex-stratified and cohort-specific models confirm the childhood poverty penalty and the protective role of education, all supporting robustness.

### Conclusion by way of informing global health policy

The childhood poverty penalty of healthy ageing, together with its strong education and wealth gradients, indicates that later-life functionings is markedly shaped by early-life material conditions and remains malleable through current socioeconomic support. National plans under the UN Decade of Healthy Ageing should embed child poverty reduction -better housing quality for example- within ageing policies, not just within child or social policies. Then, balance early- and mid-life investment. Countries with parallel HAS age profiles for poor vs non-poor should prioritise closing the initial gap (family support, early learning support, housing improvements). Where widening gaps mark the age profiles, complement early action with mid-life interventions: chronic-disease management, age-friendly environments, and financial protection to buffer the wealth penalty found here.

Further, target educational returns where they are largest. The considerable college advantage in China and Indonesia suggests that human-capital expansion can substantially raise later-life functional capacity in rapidly developing settings; conversely, sustained adult-learning programmes may help sustain healthy ageing high-income societies. On a more practical level, employ measurement more widely. Routine incorporation of life history interviews, latent indicators of childhood conditions, and metric-preserving healthy-ageing scale and its items can turn retrospective data into policy-relevant signals while avoiding unsafe inference. More ambitiously, enriching biomarker collections with methylation (as in HRS Venous blood sample I used to show that the childhood poor aged epigenetically faster)^14^ allows nations to track mechanistic links that inform prevention and rehabilitation.

Last, use such measurement and evidence to plan for long-term or social care labour. The trellis plot showing between-country levels/slopes imply very different future demands of functional support. Wealthy and developing nations should anticipate long-term care demand and develop sustainable care-worker policies (training and retention) to avoid zero-sum competition as population ageing accelerates. Healthy ageing is a life course achievement: to raise HAS fairly and efficiently, reduce childhood poverty, support older adults’ material security, and sustain functional capacity through prevention, rehabilitation and supportive environments across the arc of life.

### Ethical review

The University of Manchester exempted the investigation from full ethical review as it uses publicly available deidentified secondary datasets.

## Data Availability

All data are available from the respective repositories.

https://hrs.isr.umich.edu

https://www.elsa-project.ac.uk

https://www.share-project.org

https://tilda.tcd.ie

https://charls.pku.edu.cn

https://www.rand.org/well-being/social-and-behavioral-policy/data/FLS/IFLS.html

## Acknowledgement

I thank the participants in 32 countries around the world for providing information, time and in many visits blood biomarkers too, as well as the generous funding bodies over many decades. HRS: The HRS (Health and Retirement Study) is sponsored by the National Institute on Aging (grant number NIA U01AG009740) and is conducted by the University of Michigan. ELSA: The English Longitudinal Study of Ageing was developed by a team of researchers based at University College London, NatCen Social Research, the Institute for Fiscal Studies, the University of Manchester and the University of East Anglia. The data were collected by NatCen Social Research. The funding is currently provided by the National Institute on Aging in the US (grant numbers: 2RO1AG7644 and 2RO1AG017644-01A1), and a consortium of UK government departments coordinated by the National Institute for Health Research. SHARE: The SHARE data collection has been funded by the European Commission, DG RTD through FP5 (QLK6-CT- 2001-00360), FP6 (SHARE-I3: RII-CT-2006-062193, COMPARE: CIT5-CT-2005-028857, SHARELIFE: CIT4-CT-2006-028812), FP7 (SHARE-PREP: GA N°211909, SHARE-LEAP: GA N°227822, SHARE M4: GA N°261982, DASISH: GA N°283646) and Horizon 2020 (SHARE-DEV3: GA N°676536, SHARE-COHESION: GA N°870628, SERISS: GA N°654221, SSHOC: GA N°823782, SHARE-COVID19: GA N°101015924) and by DG Employment, Social Affairs & Inclusion through VS 2015/0195, VS 2016/0135, VS 2018/0285, VS 2019/0332, and VS 2020/0313. Additional funding from the German Ministry of Education and Research, the Max Planck Society for the Advancement of Science, the U.S. National Institute on Aging (U01_AG09740-13S2, P01_AG005842, P01_AG08291, P30_AG12815, R21_AG025169, Y1-AG-4553-01, IAG_BSR06-11, OGHA_04-064, HHSN271201300071C, RAG052527A) and from various national funding sources is gratefully acknowledged (see www.share-project.org). CHARLS: the CHARLS research and field team coordinated by Peking University. IFLS: the RAND Corporation, University of Indonesia, University of Gadjah Mada, SurveyMETER Yogyakarta.

All the data are publicly available from respective repositories (HRS) https://hrs.isr.umich.edu, (SHARE) www.share-project.org, (ELSA) www.elsa-project.ac.uk, (CHARLS) https://charls.pku.edu.cn, (IFLS) https://www.rand.org/well-being/social-and-behavioral-policy/data/FLS/IFLS.html, (TILDA) https://tilda.tcd.ie

## Acronyms

CHARLS: China Health and Retirement Longitudinal Study
ELSA: English Longitudinal Study of Ageing
HRS: Health and Retirement Study
IFLS: Indonesia Family Life Survey
NHS: National Health Service
SHARE: Survey of Health, Ageing and Retirement in Europe
TILDA: The Irish Longitudinal Study on Ageing
UN: United Nations

